# Impact of the COVID-19 Pandemic on Vaccination Coverage in Ecuador: A Pre- and Post-Pandemic Comparative Analysis

**DOI:** 10.1101/2025.03.26.25324742

**Authors:** José Daniel Sánchez, Kimberlly Pamela Montenegro Cuello, Alejandro Arjuna Rodriguez Proaño

**Affiliations:** Facultad de Ciencias de la Salud y Bienestar Humano, Universidad Indoamérica, Quito Ecuador; Facultad de Ciencias de la Salud “Eugenio Espejo”, Universidad UTE, Quito Ecuador

**Keywords:** COVID-19 pandemic, Vaccination coverage, Ecuador, Immunization, Routine vaccination, Health disparities, Vaccine hesitancy

## Abstract

This study examines the impact of the COVID-19 pandemic on vaccination coverage in Ecuador, analyzing trends before (2019) and during the pandemic (2020-2021). Using data from the Ministry of Public Health and the National Institute of Statistics and Censuses, the study compares routine childhood vaccination coverage and COVID-19 vaccination efforts. Results indicate a significant decline in routine vaccination coverage during the pandemic, with reductions observed for vaccines such as BCG, pentavalent, pneumococcal, and rotavirus. Furthermore, the study highlights socioeconomic and geographical disparities in COVID-19 vaccine coverage, with vulnerable populations facing challenges in accessing vaccination. In conclusion, the pandemic has negatively impacted routine immunization programs in Ecuador; addressing disparities in access is crucial for improving overall vaccination coverage and public health outcomes.

## 1. Introduction

The COVID-19 pandemic has precipitated a global health crisis of unprecedented scale, with profound repercussions across various facets of society. Beyond the direct morbidity and mortality inflicted by the SARS-CoV-2 virus, the pandemic has exerted a significant impact on essential healthcare services, including routine immunization programs. Globally, the diversion of resources, overburdening of health systems, and implementation of public health measures such as lockdowns and movement restrictions have led to substantial disruptions in vaccination services. These disruptions have resulted in a decline in vaccination coverage for numerous preventable diseases, posing a serious threat to global health security and potentially reversing decades of progress in disease control.

The impact of the pandemic on immunization programs has been particularly acute in low- and middle-income countries (LMICs), where healthcare infrastructure m y be fragile and resources limited. In these settings, the pandemic has not only disrupted service delivery but also exacerbated pre-existing challenges such as limited access to healthcare, supply chain vulnerabilities, and vaccine hesitancy. Consequently, the decline in vaccination coverage has increased the risk of outbreaks of vaccine-preventable diseases, including measles, poliomyelitis, diphtheria, and pertussis, further compounding the health burden faced by these nations.

Ecuador, a South American country with a diverse population and varying levels of healthcare access across its regions, has also experienced significant disruptions to its immunization programs as a result of the COVID-19 pandemic. In Ecuador, as of July 2021, only 57% of the population had received the first COVID-19 vaccine dose, highlighting significant challenges in reaching and providing vaccines to underserved populations in remote areas (1). Prior to the pandemic, Ecuador’s national immunization program faced challenges in achieving optimal coverage rates, with disparities observed across different geographical regions and socioeconomic groups. The pandemic has further exacerbated these challenges, leading to a decline in the coverage of routine childhood vaccinations and highlighting existing inequities in access to healthcare services. Specifically, the pandemic has impacted the coverage of vaccines for diseases such as measles, poliomyelitis, diphtheria, and pertussis. A study of the impact of the COVID-19 pandemic on routine vaccination coverage across 39 countries and territories in Latin America and the Caribbean (LAC) found that pre-existing declines were exacerbated. By comparing actual DTPcv (diphtheria-pertussis-tetanus-containing vaccine) first and third dose coverage with forecasted levels, the study identified significant reductions in 79% of the assessed regions. Furthermore, the study explored the influence of socioeconomic factors, immunization policies, and pandemic-related measures on these coverage changes. (2)

## 2. Materials and Methods

### Study Design

This study employed a retrospective, observational design to analyze the impact of the COVID-19 pandemic on vaccination coverage in Ecuador. The study involved a comparative analysis of vaccination coverage data for the pre-pandemic period (2019) and the pandemic period (2020-2021). This design allowed for the examination of trends and changes in vaccination coverage over time, as well as the assessment of potential associations between the pandemic and vaccination outcomes.

### Data Sources

The data for this study were obtained from the following primary sources:

- **Boletín de Indicadores de la Estrategia Nacional de Inmunización del Ministerio de Salud Pública (MSP):** This bulletin, published by the Ministry of Public Health of Ecuador, provided detailed information on routine childhood vaccination coverage at the national and sub-national levels. The data included the number of doses administered, the number of doses applied, and vaccination coverage rates for various vaccines.
- **Instituto Nacional de Estadística y Censos (INEC):** The INEC provided demographic data for Ecuador, including population estimates and birth rates, which were used to calculate vaccination coverage rates.
- **Published Literature**: To supplement the primary data sources and provide context on the broader impact of the pandemic and socioeconomic disparities, relevant peer-reviewed studies were identified through a systematic literature review. Databases such as PubMed, Scopus, and Web of Science were searched using relevant keywords.

### Study Population

The primary study population consisted of infants under one year of age and children up to 24 months of age in Ecuador. Data on routine childhood vaccinations were analyzed for this age group. Additionally, data on the general population were analyzed to assess COVID-19 vaccination coverage and disparities.

### Vaccination Coverage Metrics

The following vaccination coverage metrics were used in this study:

### Routine Childhood Vaccination Coverage

Coverage rates were calculated as the percentage of children in the target age group who had received the recommended doses of each vaccine. Specific vaccines included: BCG, pentavalent, pneumococcal, poliovirus, rotavirus, measles, yellow fever, and varicella.

### COVID-19 Vaccination Coverage: Coverage rates were assessed by

- Percentage of the population receiving at least one dose of a COVID-19 vaccine.
- Percentage of the population completing the primary vaccination series (two doses).
- Percentage of the population receiving booster doses.

### Data Analysis

Statistical analyses were performed using appropriate software. The following methods were employed:

- **Descriptive Statistics:** Descriptive statistics, including frequencies, percentages, means, and standard deviations, were used to summarize the characteristics of the study population and the vaccination coverage data.
- **Trend Analysis:** Trend analysis was used to examine changes in routine childhood vaccination coverage over the study period (2019-2021). Coverage rates were plotted over time to visualize trends and identify significant changes.
- **Comparative Analysis:** Vaccination coverage rates were compared between the pre-pandemic year (2019) and the pandemic years (2020-2021) to assess the impact of the pandemic on vaccination coverage.

A comparative analysis of vaccine doses administered from 2018 to 2020 revealed a significant decline in 2020, with a reduction of approximately 137,000 doses compared to the stable rates of the previous two years. This decrease was reflected in coverage percentages, with the pentavalent vaccine showing the most substantial drop (17.7%), followed by poliovirus, rotavirus, and pneumococcal vaccines. Spatial analysis indicated that coastal and highland provinces experienced the most severe reductions in vaccination coverage, highlighting regional disparities in the pandemic’s impact on immunization services.(3)

## 3. Results

Analysis of routine childhood vaccination data revealed a clear trend of decreasing vaccination coverage between 2019 and 2021, demonstrating a significant impact of the COVID-19 pandemic on adherence to immunization schedules. In 2019, the majority of vaccines demonstrated coverage rates exceeding 80%; however, these rates experienced a substantial decline in both 2020 and 2021. Specifically, the coverage of the BCG vaccine, administered at birth, decreased from 86.5% in 2019 to 80.6% in 2020, and further to 75.3% in 2021. This pattern of reduction was also observed across other essential vaccines, including pentavalent, pneumococcal, and rotavirus vaccines, all of which showed a significant decrease in administration. While the decline in coverage was already noticeable in 2020, 2021 was marked by an even more pronounced reduction, with coverage rates for some vaccines falling below 60%. For instance, coverage rates for vaccines such as SRP 2 (which protects against measles, rubella, and mumps and is administered at 18 months) and bOPV (the bivalent oral poliovirus vaccine administered at 12 months) were 75.7% and 76.7%, respectively, in 2019. By 2020, these rates had decreased to 68.0%, and by 2021, they had fallen further to 58.0%. This reduction in vaccination coverage has important implications for public health, as it can elevate the risk of outbreaks of vaccine-preventable diseases within the pediatric population.

WHO has proposed the development of combined vaccines because of their advantages; in Latin America certain projects were started to this end, although the development of these vaccines is complex due to their technological challenges because of the possible negative interactions between the antigens. The National Institute of Hygiene of Ecuador asked for the collaboration of the CIGB to develop a pentavalent vaccine (DPT-HB-Hib) in Ecuador with diphtherial and tetanus anatoxin, whole cells of Bordetella pertussis (produced in Ecuador), recombinant hepatitis B virus surface antigen and the synthetic polysaccharide (polirribosyl ribitol phosphate, PRP) of Haemophilus influenzae conjugated to tetanus anatoxin (PRP-T) produced at the CIGB.(4)

Table 2 presents the number of doses administered, the type of vaccine applied, and the vaccination coverage among children of different age groups (from newborns to early childhood). A progressive decline in vaccination coverage between 2019 and 2021 is observed, suggesting a significant impact of the COVID-19 pandemic on adherence to the immunization schedule.

**Table 1.** Region and Province Data for Years 2019, 2020, and 2021.

| Region | Province | 2019 | 2020 | 2021 |
| --- | --- | --- | --- | --- |
| Costa | Esmeraldas | 13.293 | 13.211 | 13.128 |
|  | Manabi | 29.299 | 29.207 | 29.005 |
|  | Los Rios | 18.897 | 18.888 | 18.798 |
|  | Santa Elena | 8.834 | 8.897 | 8.900 |
|  | Guayas | 79.543 | 79.535 | 79.519 |
|  | Santo Domingo | 10.535 | 10.537 | 10.541 |
|  | El Oro | 12.526 | 12.464 | 12.438 |
| Sierra | Azuay | 15.903 | 15.7 | 15.688 |
|  | Bolivar | 4.338 | 4.223 | 4.205 |
|  | Cañar | 5.680 | 5.660 | 5.640 |
|  | Carchi | 3.258 | 3.236 | 3.214 |
|  | Cotopaxi | 10.355 | 10.304 | 10.293 |
|  | Chimborazo | 9.853 | 9.764 | 9.660 |
|  | Imbabura | 9.173 | 9.141 | 9.115 |
|  | Loja | 9.978 | 9.923 | 9.872 |
|  | Pichincha | 56.698 | 57.062 | 57.200 |
|  | Tungurahua | 10.166 | 10.111 | 10.069 |
| Amazonia | Morona Santiago | 4.895 | 4.842 | 4.822 |
|  | Napo | 3.341 | 3.361 | 3.381 |
|  | Orellana | 3.883 | 3.821 | 3.800 |
|  | Pastaza | 2.639 | 2.659 | 2.679 |
|  | Sucumbios | 4.944 | 4.958 | 4.978 |
|  | Zamora Chinchipe | 2.839 | 2.837 | 2.833 |
| Insular | Galápagos | 624 | 631 | 666 |
|  | Total: | 331.494 | 330.972 | 330.444 |

<sup>1</sup> DNEAIS - Early and late capture base. Developed by: MSP national immunization strategy.

**Table 2.**
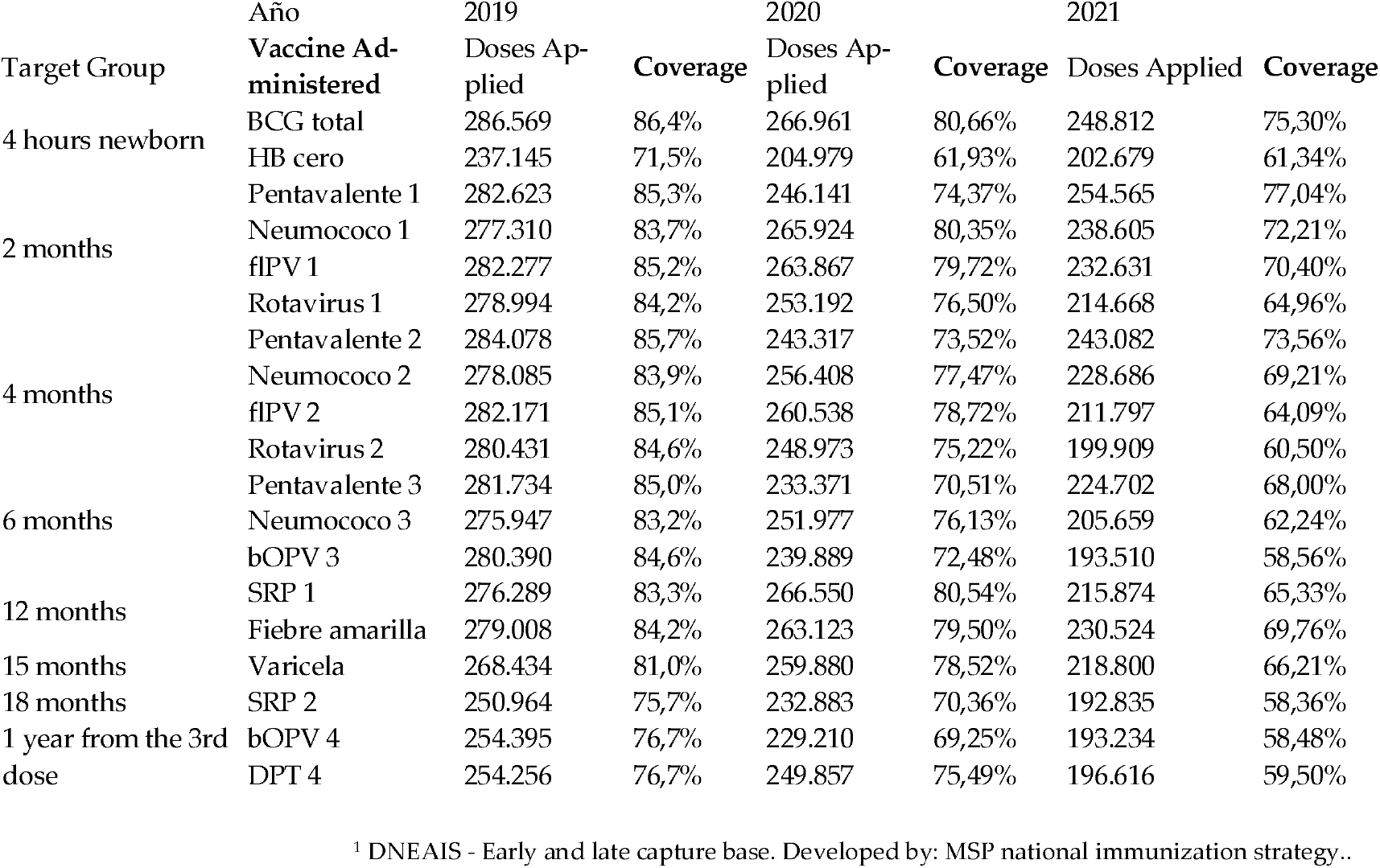
Coverage Years 2019, 2020, and 2021.

### General Decline in Vaccination Coverage

In 2019, most vaccines had coverage rates above 80%; whereas in 2020 and 2021, these figures dropped considerably. For the BCG vaccine (administered at birth), coverage decreased from 86.5% in 2019 to 80.6% in 2020, and further to 75.3% in 2021. A similar pattern is observed for other essential vaccines, such as pentavalent, pneumococcal, and rotavirus, all showing significant reductions in administration.

### Greater Impact in 2021

Although the decline in coverage was already evident in 2020, the year 2021 shows an even more marked reduction, with coverage rates in some cases falling below 60%. Vaccines such as MMR 2 (measles, mumps, and rubella) administered at 18 months and bOPV at 12 months had coverage rates of 75.7% and 76.7% in 2019, respectively, decreasing to 68.0% in 2020 and dropping further to 58.0% in 2021.

### Public Health Implications

The reduction in vaccination coverage can increase the risk of outbreaks of preventable diseases among the pediatric population. It is crucial to reinforce and reorganize strategies to recover coverage rates through mass campaigns (as was done in 2023), follow-up with unvaccinated children, and improved access to health services for the entire population. The COVID-19 pandemic significantly disrupted infant immunization programs across Ecuador, underscoring the urgent need for targeted interventions to safeguard vulnerable populations during public health emergencies.(3)

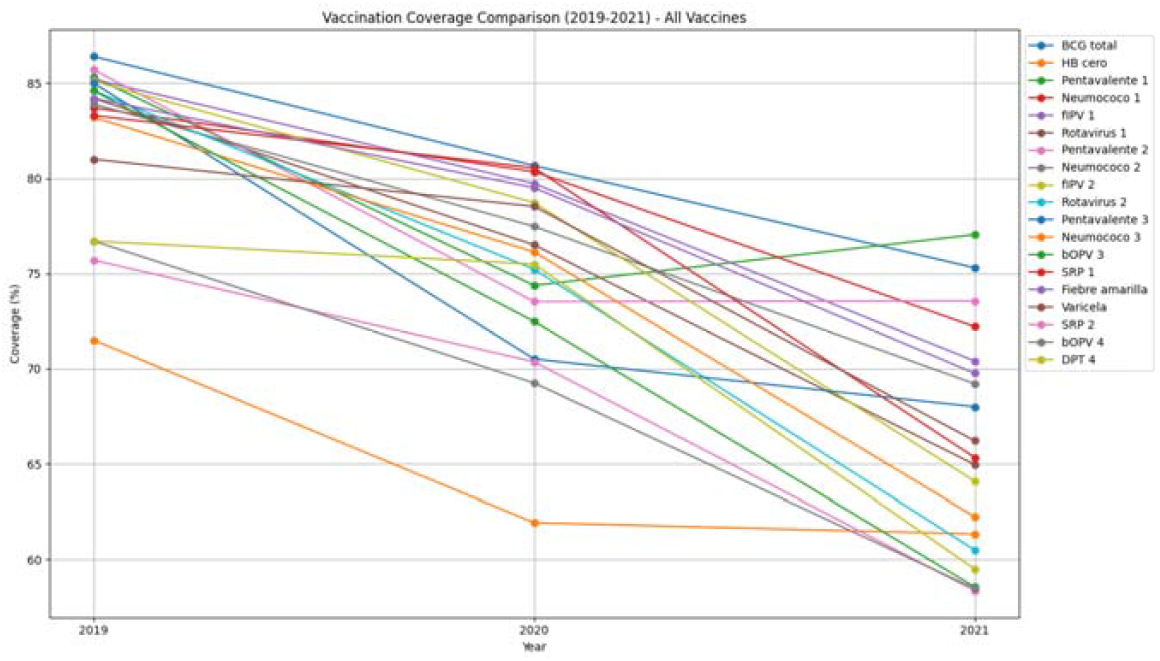

The SARS-CoV-2 pandemic significantly disrupted critical health programs, including childhood immunization. To identify areas at risk for communicable diseases and analyze vaccination coverage in rural and urban parishes within the Metropolitan District of Quito from 2019 to 2022, an exploratory ecological observational study was conducted. Consolidated data were utilized to calculate coverage indicators for both urban and rural parishes. Temporal trends in monthly vaccination coverage were assessed using Joinpoint regression analysis, and risk areas were visualized through mapping.

Results indicated that in urban parishes, pentavalent vaccine coverage increased from 70% in 2019 to 96% in 2022. Conversely, rural parishes experienced a marked decline, with coverage rates decreasing from 42% in 2019 to 66% in 2022. The rural parish of Zámbiza showed the highest coverage, while Chavezpamba had the lowest. In urban areas, Centro Histórico had the highest coverage, and Iñaquito the lowest. Joinpoint analysis identified a significant drop in vaccination coverage in April 2020, coinciding with the peak of the COVID-19 pandemic. (5)

## 4. Discussion

The findings of this study reveal a concerning trend of declining vaccination coverage in Ecuador during the COVID-19 pandemic. The data clearly illustrate a disruption in routine immunization programs, with a decrease in the coverage of key childhood vaccines. This decline has significant implications for public health and highlights the far-reaching consequences of the pandemic beyond the direct impact of the virus itself. The observed reduction in vaccination coverage aligns with global trends. The pandemic strained response efforts. In Ecuador, this strain manifested as a change in health services and the overwhelming of hospitals and health centers, as detailed in the introduction. Consequently, routine immunization programs were disrupted, leading to decreased vaccination rates. Several factors contributed to this decline: movement restrictions and lock-downs, implemented to curb the spread of the virus, limited access to healthcare facilities. As noted in the introduction, increased fear of contagion also deterred families from visiting health centers for vaccinations. Logistical challenges, including disruptions in vaccine supply chains, further compounded the problem. These challenges collectively created barriers to vaccination, resulting in decreased coverage across various vaccines. The decline in coverage for vaccines like BCG, pentavalent, pneumococcal, and rotavirus is particularly concerning. These vaccines protect against serious childhood diseases, and a decrease in their coverage increases the risk of outbreaks. As highlighted in the introduction, the low vaccination rates have increased the possibility of the resurgence of diseases such as rubella and measles, with suspected cases already being reported in some parts of the country. Even for diseases like polio, where Ecuador has maintained a polio-free status since 1990, the reduced OPV vaccination rate has raised public health concerns. The need to reinforce and reorganize vaccination strategies is critical. As the Results section notes, strategies such as mass campaigns, follow-up with unvaccinated children, and improving access to health services for the entire population are essential to recover vaccination coverage. The long-term consequences of the pandemic on immunization programs necessitate proactive measures to mitigate their negative effects and build more resilient immunization systems. While this study provides valuable insights into the pandemic’s impact on vaccination coverage in Ecuador, it is important to acknowledge its limitations. First, the analysis does not fully capture the impact of socioeconomic determinants on vaccination coverage. Second, detailed data at the provincial level were not consistently available for all vaccines and years, limiting the granularity of geographical comparisons. Despite these limitations, the study’s findings underscore the urgent need to address the decline in vaccination coverage in Ecuador.

## 5. Conclusions

The evidence presented demonstrates the profound impact of the COVID-19 pandemic on vaccination coverage in Ecuador. The global health crisis triggered a cascade of disruptions that reverberated through the nation’s immunization programs, leading to a notable decline in the uptake of routine childhood vaccinations. This decline is particularly concerning given the potential resurgence of vaccine-preventable diseases, which pose a significant threat to public health, especially among the most vulnerable populations. The data reveal a clear deterioration in coverage rates for critical vaccines, including BCG, pentavalent, pneumococcal, and rotavirus. This reduction not only reflects the immediate challenges posed by the pandemic, such as healthcare service disruptions and mobility restrictions, but also underscores pre-existing vulnerabilities within the Ecuadorian health system. Factors like limited access to healthcare, supply chain issues, and vaccine hesitancy were likely exacerbated by the pandemic, creating a complex web of obstacles to effective immunization. While the immediate focus was understandably on combating COVID-19, the collateral damage to routine immunization programs necessitates urgent attention. The findings emphasize the need for robust recovery strategies to restore and enhance vaccination coverage. These strategies must address both the systemic challenges within the healthcare infrastructure and the socio-behavioral factors influencing vaccine uptake. Looking ahead, it is crucial for Ecuador to prioritize the strengthening of its immunization systems to build resilience against future health crises. This involves not only ensuring adequate resources and infrastructure but also implementing targeted interventions to reach underserved populations, combat vaccine misinformation, and promote vaccine confidence. Continued monitoring, evaluation, and research are essential to track progress, identify emerging challenges, and adapt strategies to achieve optimal vaccination coverage and safeguard public health gains. In summary, the COVID-19 pandemic has presented Ecuador with a significant setback in its efforts to maintain and improve vaccination coverage. Overcoming the challenges and mitigating the long-term consequences will require sustained commitment, strategic investment, and collaborative action across all levels of the healthcare system and society.

## Data Availability

Data Availability Statement: All data used in this study were obtained from publicly available sources including the Boletin de Indicadores de la Estrategia Nacional de Inmunizacion del Ministerio de Salud Publica (MSP) and the Instituto Nacional de Estadistica y Censos (INEC) of Ecuador. The datasets analyzed during the current study are available from the corresponding author upon reasonable request.

## Author Contributions

Kimberlly Pamela Montenegro Cuello and Alejandro Arjuna Rodriguez Proaño contributed to the validation, formal analysis, and statistical analysis; José Daniel Sánchez, principal co-author, contributed to the conceptualization, methodology, investigation, resource acquisition, data curation, original draft preparation, manuscript review and editing, visualization, supervision, and project administration.

## Funding

This research received no external funding. The APC was funded by Universidad Indoamérica.

## Institutional Review Board Statement

Not applicable. This study did not involve human or animal subjects, and therefore did not require ethical review or approval from an Institutional Review Board (IRB).

## Informed Consent Statement

Not applicable. This study utilized secondary, publicly available external databases that are freely accessible and do not require individual participant consent. The research was conducted using anonymized, aggregate data sources that are in the public domain, and no individual human subjects were directly involved in the research process.

## Abbreviations

The following abbreviations are used in this manuscript:

MDPI: Multidisciplinary Digital Publishing Institute
DOAJ: Directory of Open Access Journals
TLA: Three Letter Acronym
LD: Linear Dichroism
LMICs: Low- and Middle-Income Countries
MSP: Ministry of Public Health
INEC: National Institute of Statistics and Censuses
BCG: Bacillus Calmette–Guérin (vaccine)
HB: Hepatitis B
flPV: Inactivated Polio Vaccine
bOPV: Bivalent Oral Poliovirus
SRP: Measles, Rubella, and Mumps
DPT: Diphtheria, Pertussis, and Tetanus
MMR: Measles, Mumps, and Rubella

## Disclaimer/Publisher’s Note

The statements, opinions and data contained in all publications are solely those of the individual author(s) and contributor(s) and not of MDPI and/or the editor(s). MDPI and/or the editor(s) disclaim responsibility for any injury to people or property resulting from any ideas, methods, instructions or products referred to in the content.

## Notes

### Competing Interest Statement

The authors have declared no competing interest.

### Funding Statement

Funding: This research received no external funding. The Article Processing Charge (APC) was funded by Universidad Indoamerica. Neither the authors nor their institutions received payment or services from any third party for any aspect of the submitted work, including grants, data monitoring board, study design, manuscript preparation, or statistical analysis. The authors declare that no financial relationships existed with entities that could have influenced the submitted work

